# Empirical Assessment of COVID-19 Crisis Standards of Care Guidelines

**DOI:** 10.1101/2020.05.16.20098657

**Authors:** Julia L. Jezmir, Maheetha Bharadwaj, Sandeep P. Kishore, Marisa Winkler, Bradford Diephuis, Edy Y. Kim, William B. Feldman

**Affiliations:** Department of Medicine, Brigham and Women’s Hospital, Boston, MA 02115; Harvard Medical School, Boston, MA 02115; Division of Pulmonary and Critical Care Medicine, Department of Medicine, Brigham and Women’s Hospital, Boston, MA 02115

**Author notes:** Indicates equal contributions as co-first authors (Julia L. Jezmir, Maheetha Bharadwaj, Sandeep P. Kishore). Indicates equal contributions as co-senior authors (Edy Y. Kim, William B. Feldman). Co-corresponding authors. **Corresponding authors:** William B. Feldman, MD, DPhil, Division of Pulmonary and Critical Care Medicine, Department of Medicine, Brigham and Women’s Hospital, Boston, MA 02115, Edy Y. Kim, MD, PhD, Division of Pulmonary and Critical Care Medicine, Department of Medicine, Brigham and Women’s Hospital, Boston, MA 02115.

## Abstract

**Background:** Several states have released Crisis Standards of Care (CSC) guidelines for the allocation of scarce critical care resources. Most guidelines rely on Sequential Organ Failure Assessment (SOFA) scores to maximize lives saved, but states have adopted different stances on whether to maximize long-term outcomes (life-years saved) by accounting for patient comorbidities.

**Methods:** We compared 4 representative state guidelines with varying approaches to comorbidities and analyzed how CSC prioritization correlates with clinical outcomes. We included 27 laboratory-confirmed COVID-19 patients admitted to ICUs at Brigham and Women’s Hospital from March 12 to April 3, 2020. We compared prioritization algorithms from New York, which assigns priority based on SOFA alone; Maryland, which uses SOFA plus severe comorbidities; Pennsylvania, which uses SOFA plus major and severe comorbidities; and Colorado, which uses SOFA plus a modified Charlson comorbidity index.

**Results:** In pairwise comparisons across all possible pairs, we found that state guidelines frequently resulted in tie-breakers based on age or lottery: New York 100% of the time (100% resolved by lottery), Pennsylvania 86% of the time (18% by lottery), Maryland 93% of the time (35% by lottery), and Colorado: 32% of the time (10% by lottery). The prioritization algorithm with the strongest correlation with 14-day outcomes was Colorado (r_s_ = −0.483. p = 0.011) followed by Maryland (r_s_ = −0. 394, p =0.042), Pennsylvania (r_s_ = −0.382, p = 0.049), and New York (r_s_ = 0). An alternative model using raw SOFA scores alone was moderately correlated with outcomes (r_s_ = −0.448, p = 0.019).

**Conclusions:** State guidelines for scarce resource allocation frequently resulted in identical priority scores, requiring tie-breakers based on age or lottery. These findings suggest that state CSC guidelines should be further assessed empirically to understand whether they meet their goals.

## Background

The COVID-19 pandemic has renewed debate about crisis standards of care (CSC).^1,2^ Several states have released guidelines for the allocation of critical care resources, including ventilators, should they become scarce.^3^ Most guidelines rely upon Sequential Organ Failure Assessment (SOFA) scores to identify those likely to survive hospitalization and thereby maximize the number of lives-saved.^3^ However, states have adopted differing stances on whether to maximize long-term outcomes (i.e. life-years saved) by accounting for patient comorbidities.

We compared four state guidelines with varying approaches to patient comorbidities: New York, which assigns priority scores based on SOFA alone; Maryland, which uses SOFA plus severe comorbidities (< 1 year expected survival); Pennsylvania, which uses SOFA plus major and severe comorbidities; and Colorado, which uses SOFA plus a modified Charlson comorbidity index (eTables 1-2).^3,4^ We assessed whether these guidelines produce a range of priority scores (or instead required tie-breakers) and whether prioritization is associated with 14-day outcomes.

## Methods

We included patients with laboratory-confirmed COVID-19 admitted to ICUs at Brigham and Women’s Hospital from March 12 to April 3, 2020. We followed each state guideline when calculating SOFA category (none used raw SOFA score) and breaking ties between patients with the same priority scores. All states relied on lotteries to break ties, but Pennsylvania, Maryland, and Colorado used age-categories first before random allocation. Scores were calculated immediately preceding intubation to model real-world triage. Outcomes were based on a 7-point scale ranging from death (1) to hospital discharge (7).^5^ Two independent reviewers manually abstracted data from electronic health records; conflicts were adjudicated by a third reviewer. Correlations were calculated using the Spearman’s rank correlation coefficient, accounting for tie-breakers when indicated. Statistical tests were performed in IBM SPSS Statistics Version 25.0. Approval was obtained from the Partners HealthCare Institutional Review Board.

## Results

Twenty-seven patients met criteria for inclusion (eTable 3). At 14 days of follow-up, 7 patients (26%) had died, 8 (30%) were on mechanical ventilators, 8 (30%) were hospitalized without requiring supplemental oxygen, and 4 (15%) had been discharged. In pairwise comparisons of all possible patient pairs, tie-breakers were needed 100% of the time by New York guidelines (100% of which were resolved by lottery), 86% of the time by Pennsylvania (18% by lottery), 93% of the time by Maryland (35% by lottery), and 32% of the time by Colorado (10% by lottery) (Table 1). Similar correlations were observed between priority assignments and 14-day outcomes in Colorado (r_s_ = −0.483. p = 0.011), Maryland (r_s_ = −0.394, p =0.042), and Pennsylvania (r_s_ = −0.382, p = 0.049) (Table 2). No correlation was observed in New York (r_s_ = 0), though we did observe correlation when using raw SOFA scores (r_s_ = −0.448, p = 0.019).

**Table 1:** CSC Guidelines Performance Across Pairwise Comparisons.

Pairwise comparisons were conducted for all possible pairs for each of the State Guidelines and Alternative Models to assess how well each algorithm performed in selecting patients with better outcomes at 14 days from intubation.
|  | State Guidelines |  |  |  | Alternative Models |  |
| --- | --- | --- | --- | --- | --- | --- |
|  | Colorado Guidelines | Maryland Guidelines | Pennsylvania Guidelines | New York Guidelines | SOFA Score | Age |
| **Algorithm gave higher priority to the patient with the:** |  |  |  |  |  |  |
| Better outcome | 52% | 38% | 45% | 0% | 46% | 51% |
| Worse outcome | 19% | 13% | 19% | 0% | 18% | 26% |
| Equal outcome | 19% | 15% | 18% | 0% | 17% | 21% |
| **Algorithm gave equal priority to both patients:** |  |  |  |  |  |  |
| Requires lottery | 10% | 35% | 18% | 100% | 19% | 2% |

**Table 2:** Correlations of CSC Priority Scoring Guidelines and Outcomes.

| CSC Priority Scoring Algorithms | Spearman's Correlation ($\rho$) | p-value |
| --- | --- | --- |
| **State Guidelines<sup>a</sup>** |  |  |
| Colorado | -0.483 | 0.011 |
| Maryland | -0.394 | 0.042 |
| Pennsylvania | -0.382 | 0.049 |
| New York<sup>b</sup> | n/a | n/a |
| **Alternative Models<sup>c</sup>** |  |  |
| SOFA Scores | -0.448 | 0.019 |
| Age | -0.374 | 0.055 |

<sup>a</sup>Correlations for state guidelines account for priority scores and tie breakers when indicated.
<sup>b</sup>NY guidelines for SOFA score calculations do not include mechanical ventilation in the P:F ratio score definition.
<sup>c</sup>SOFA scores and age were poorly correlated ( $r_s = 0.123$ , $p = 0.543$ ).

## Discussion

State guidelines for scarce resource allocation frequently resulted in identical priority scores, requiring tie-breakers based on age or lottery. Priority scores were weakly or moderately correlated with 14-day outcomes for all states except New York. Colorado guidelines, which utilized a comorbidity index, had the fewest tie-breakers and strongest correlation with 14-day outcomes.

One worry with incorporating comorbidities into triage algorithms is the risk of exacerbating racial and socioeconomic disparities.^6^ That state guidelines commonly result in tie-breakers hinging on age or lottery may help allay these concerns, though it certainly does not resolve them. Our findings raise further worries about ageism and the arbitrary nature of lotteries. One potential solution may be to utilize raw SOFA scores alone (which were moderately correlated with 14-day outcomes) and exclude comorbidities, but this may place undue emphasis on a tool that was never designed for triage.

Our study is limited by small sample size and inclusion of only COVID-19 patients. A key question for future research is how SOFA and comorbidity scores correlate with long-term outcomes. However, we have identified important limitations of several representative state triage algorithms and provided a framework for future quantitative analyses. The ethical defensibility of these algorithms will depend, in part, on empirical analyses of how they function in practice.

## Data Availability

Data are available upon request.

